# The Impact of Pre-exposure Prophylaxis for Human Immunodeficiency Virus on Gonorrhea Prevalence

**DOI:** 10.1101/19005207

**Authors:** Joe Pharaon, Chris T. Bauch

## Abstract

Pre-exposure prophylaxis (PrEP) has been shown to be highly effective in reducing the risk of HIV infection in gay and bisexual men who have sex with men (GbMSM). However, PrEP does not protect against other sexually transmitted infections (STIs). In some populations, PrEP has also led to riskier behaviour such as reduced condom usage, with the result that the prevalence of bacterial STIs like gonorrhea has increased. Here we develop a compartmental model of the transmission of HIV and gonorrhea, and the impacts of PrEP, condom usage, STI testing frequency and potential changes in sexual risk behaviour stemming from the introduction of PrEP in a population of GbMSM. We find that introducing PrEP causes an increase in gonorrhea prevalence for a wide range of parameter values, including at the current recommended frequency of STI testing once every 3 months for individuals on PrEP. Moreover, the model predicts that a higher STI testing frequency alone is not enough to prevent a rise in gonorrhea prevalence, unless the testing frequency is increased to impractical levels. However, testing every 2 months in combination with sufficiently high condom usage by individuals on PrEP would be successful in maintaining gonorrhea prevalence at pre-PrEP levels. The results emphasize that programs making PrEP more available should be accompanied by efforts to support condom usage and frequent STI testing, in order to avoid an increase in the prevalence of gonorrhea and other bacterial STIs.

## 1 Introduction

Sexually transmitted infections have long been a public health concern in the GbMSM (gay and bisexual men who have sex with men) community, especially since the sexual liberation movement and the emergence of sex clubs and bath houses in the late 1970s [1, 2]. At around the same time, the Human Immunodeficiency Virus, known as HIV, started spreading and the first cases were recorded. HIV patients progressed to AIDS (Acquired Immunodeficiency Syndrome) and subsequently died. An AIDS pandemic began in the 1980s. The origins of HIV are still debated in the scientific community and multiple theories have been laid out [3, 4]. Nevertheless, it is a virus that disproportionately affects sexually active gay and bisexual men in Canada and North America.

HIV has two main types (1 and 2) and several subtypes [4]. The strain that affects most gay and bisexual men is type 1 subtype B [5]. HIV is a retrovirus–a class of viruses that quickly mutate–thus making it difficult to cure with antiviral drugs [6]. As the pandemic accelerated, research began to understand the modes of transmission. HIV virus particles (known as virions) are mainly found in pre-ejaculatory fluid and semen and are transmitted through anal intercourse and oral sex [7]. For a long time, condoms have been identified as the main tool for HIV prevention. Condoms lower the transmission risk of HIV by 60% to 90% [8, 9].

Testing for HIV became available in 1985 [10]. Public health agencies and physicians recommend frequent testing, especially for highly sexually active individuals. If an individual tests positive for HIV, they initiate ART (Anti Retroviral Therapy) or HAART (Highly Active Anti Retroviral Therapy) treatment which consists in taking daily pills (Truvada in combination with other medications) to bring viral loads to undetectable levels: lower than 50 copies of virus particles per milliliters of blood. Once the patient achieves the undetectable status, they can no longer transmit the virus [11].

Other sexually transmitted infections (STIs) such as gonorrhea, syphilis and chlamydia are also mainly transmitted through anal intercourse and oral sex. The three aforementioned diseases are bacterial and are usually treated with antibiotics [12]. However, if they are not diagnosed in their early stages, they can lead to complications resulting in severe health issues [13]. Condoms also act as a prophylactic measure against them. Individuals can be co-infected by HIV and other STIs [14]. If an individual is already infected by one STI, it is easier to contract another STI. gonorrhea is becoming resistant to antibiotics [13] and drug-resistant gonorrhea is becoming a public health concern.

In recent years, a treatment protocol based on antiviral drugs called pre-exposure prophylaxis (PrEP) has been implemented, and has been shown to strongly reduce the transmission of HIV [15, 16]. Recent studies have shown that PrEP can protect well above 90% [17]. However, PrEP doesn’t protect against other STIs, such as gonorrhea. Studies have associated the use of PrEP with an increase in risky sexual behaviour (such as reduced condom usage) [18, 19, 20] as well as an increase in bacterial STIs including gonorrhea [21]. Physicians, nurse practitioners and public health agents prescribe PrEP for 3 months and require the user to return with negative results for an inclusive STI testing for a refill.

PrEP is currently in the market and is being prescribed to individuals deemed at risk determined by their sex practices. It is covered by most health insurance companies and provincial and federal governments [22]. A very high PrEP uptake could potentially result in larger than average outbreaks in gonorrhea.

HIV transmission and the impact of interventions such as antiviral drugs, condoms, and hypothetical vaccines have been a frequent topic of mathematical modelling efforts [23, 24, 25, 26, 27, 28, 29, 30]. A few of these models have also considered interactions between HIV and other infections, such as the effects of tuberculosis coinfection with HIV [23]. Mathematical models can be useful for anticipating undesirable dynamics that emerge in the wake of infectious disease interventions, and how to best counter them, such as exemplified by increases in congenital rubella syndrome incidence under certain rubella vaccination policies [31]. The observed increase in gonorrhea due to use of PrEP in some populations is another example of an undesirable side effect of increased PrEP uptake. Here, we develop a model of the HIV and gonorrhea transmission in a population of GbMSM, including STI testing, condom usage, and PrEP adoption. The model is parameterized with data from Canadian and United States GbMSM populations. We use the model to study the relationship between STI testing frequency and gonorrhea prevalence after the introduction of PrEP, with a particular focus on how much STI testing frequency needs to be increased in order to counter a potential rise in gohorrhea due to reduced condom usage among individuals taking PrEP. The model is described in the following section, followed by the results and finally a discussion section.

## 2 Methods

### 2.1 Model

We introduce a system of ordinary differential equations that describes the spread of human immunodeficiency virus (HIV) and gonorrhea (GC) in the presence of a pre-exposure prophylaxis (PrEP) regimen in a population of gay and bisexual men. The population is divided into eight compartments and each individual can belong to only one compartment at a time. The compartments and the possible transfers between them appear in Figure 1.

**Figure 1:**
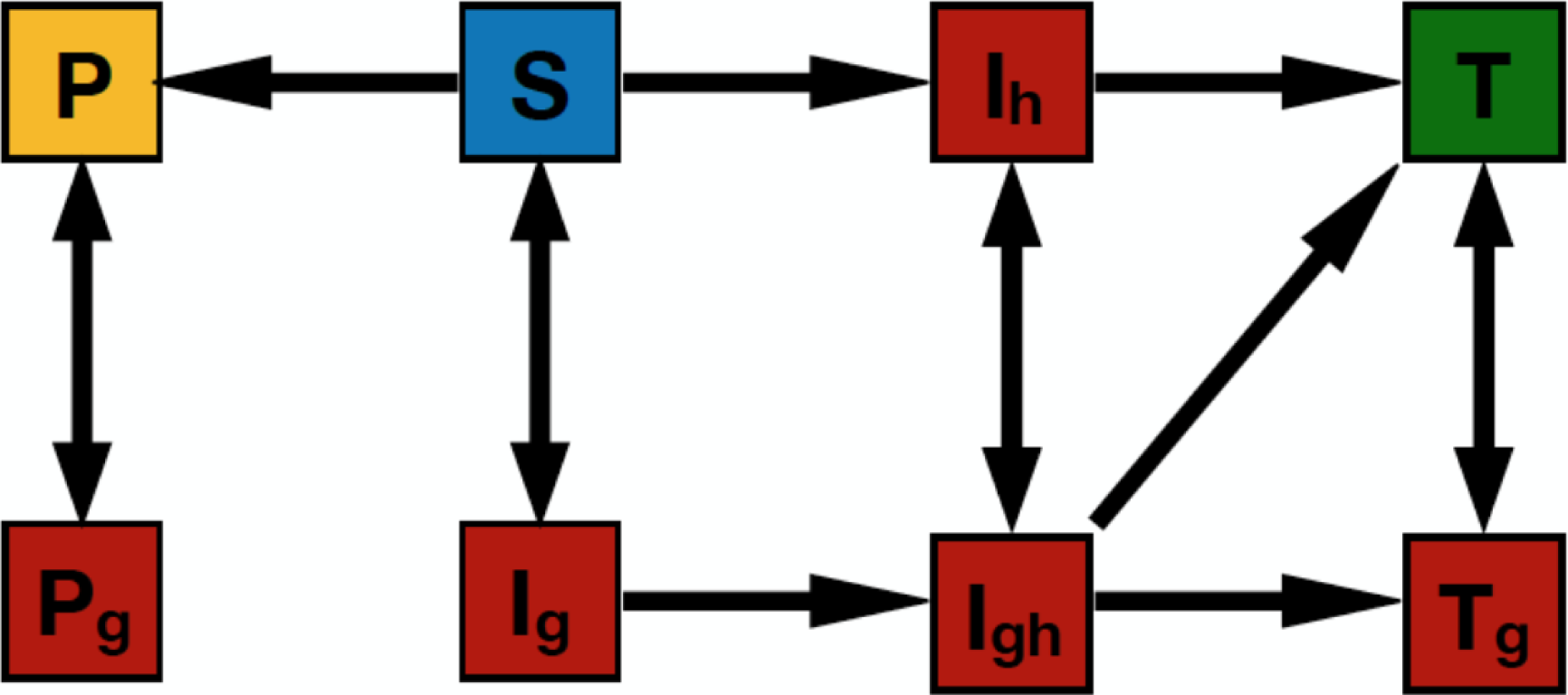
Diagram representing model structure. The eight compartments represent possible infection and treatment status for HIV and gonorrhea, while arrows represent transfers between those states. See main text and Table 1 for definitions of variables and parameters.

*S*(*t*) represents the proportion of individuals who are susceptible to HIV and GC infection but are not on PrEP. *P* (*t*) represents the proportion of individuals who are both susceptible to HIV and GC infection and are also on PrEP. We assume that PrEP users strongly adhere to the regimen recommended by their physicians. The most popular recommended dosage is a daily pill of Tenofovir(TDF)/Emtricitabine(FTC). Studies have shown that strong adherence reduces the transmission of HIV by 94% [17, 32]. A more recent study of HIV transmission among serodiscordant couples has shown 0 cases of transmission when PrEP was administered as recommended [33]. We therefore assume that strong adherence prevents the transmission of HIV. We furthermore assume that PrEP patients stay on PrEP and do not discontinue its use.

**Table 1:** Baseline parameter values. The assumed values of *δ* and *µ* are based on an average lifespan of 50 years. The parameters *β*_*h*_ and *β*_*g*_ and *α*_*t*_ were first calibrated neglecting all parameters and variables related to PrEP. Finally, *α*_*p*_ and *ρ* were calibrated.

| Parameter | Description | Value (per day) | Reference |
| --- | --- | --- | --- |
| $\beta_h$ | Transmission rate of HIV | 0.001275 | Calibrated |
| $\beta_g$ | Transmission rate of GC | 0.001475 | Calibrated |
| $\alpha_p$ | GC transmission risk factor for PrEP users | 12.75 | Calibrated |
| $\alpha_t$ | GC transmission risk factor for HART patients | 3.875 | Calibrated |
| $\beta_{hg}$ | Transmission rate of GC with HIV infection | $3\beta_g$ | [62] |
| $\beta_{gh}$ | Transmission rate of HIV with GC infection | $3\beta_h$ | [62] |
| $\rho$ | PrEP uptake rate | 0.000002375 | Calibrated |
| $\delta$ | Natural death rate | 1/20075 | Assumed |
| $\mu$ | Migration rate | 1/20075 | Assumed |
| $f$ | Testing frequency for HIV/GC | 0.54/365 | [63] |
| $f_p$ | Testing frequency for HIV/GC for PrEP users | 1/90 | [44] |
| $f_t$ | Testing frequency for HIV/GC for HART patients | 1/180 | Assumed |
| $\epsilon_h$ | Efficacy of HART treatment | 0.9 (unitless) | [64] |
| $\epsilon_g$ | Efficacy of GC treatment | 0.83 (unitless) | [65, 66] |

*P*_*g*_(*t*) is the proportion of individuals who are infected by gonorrhea while on PrEP. They can recover when they test positive for gonorrhea and start medication with efficacy *ϵ*_*g*_. *I*_*g*_(*t*) represents the proportion of individuals who are infected by gonorrhea only. *I*_*h*_(*t*) is the proportion of individuals who are infected by HIV only. *I*_*gh*_(*t*) represents the proportion of individuals who are infected by both HIV and gonorrhea. Even though simultaneous co-infection is a possible outcome of a sexual act, we will neglect this outcome and assume that individuals are first infected by one pathogen, and then independently infected by the other.

*T* (*t*) represents the proportion of individuals who are on treatment for HIV infection. Individuals who are on an HIV treatment protocol do not transmit HIV [11]. They can, however, contract gonorrhea and move to the *T*_*g*_(*t*) class. We assume that individuals infected by HIV do not progress to AIDS due to the availability of HIV medication provided by governments and the fact that it can take up to 10 years for HIV to progress into AIDS [34]. The system of differential equations describing the transfers between these eight compartments are:

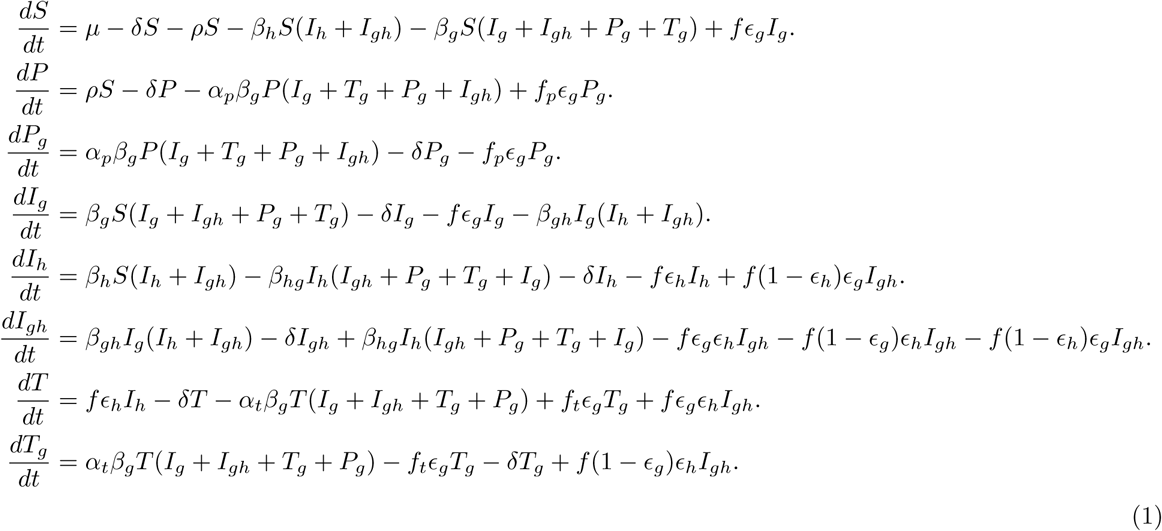

where the parameter values are defined in Table 1.

### 2.2 Parameterization

Five parameters (*ρ,α*_*p*_,*α*_*t*_,*β*_*g*_ and *β*_*h*_) were fitted to empirical values of disease prevalence to generate the baseline parameter values listed in Table 1 (all parameters that represent time rates are in units of per day, unless otherwise specified). The calibration process was carried out in two steps. The first step sets *ρ* = *α*_*p*_ = 0 and *P* (0) = *P*_*g*_(0) = 0. HIV prevalence in Canada among men who have sex with men (MSM) is roughly 19%. This number was obtained after weighing the prevalence of HIV in the main 3 metropolitan cities in Canada: Toronto, Vancouver and Montreal [35, 36]. Therefore, at equilibrium, we set *I*_*h*_ + *I*_*gh*_ + *T* + *T*_*g*_ = 0.19. The same studies reported that roughly two-thirds of HIV positive individuals are on anti-retroviral therapy (ART) or highly active anti-retroviral therapy (HAART) and therefore we set *I*_*h*_ + *I*_*gh*_ = 0.06 and *T* + *T*_*g*_ = 0.13.

Gonorrhea prevalence in MSM populations was more difficult to obtain, mostly due to disease surveillance being performed at the level of the general population (i.e., not specific to MSM). However a clinical study in California [37] reported that 15% of patients visiting a sexual health clinic were infected by gonorrhea. At equilibrium, we set *I*_*g*_ + *I*_*gh*_ + *T*_*g*_ = 0.15. The study also filters gonorrhea infection by HIV status. Taking into account only HIV negative individuals (we discarded unknown HIV status), we obtained, at equilibrium *I*_*g*_ = 0.10. We are left with *I*_*gh*_ + *T*_*g*_ = 0.05. Finally, at equilibrium *S* = 1 − 0.19 − 0.1 = 0.71.

We performed a three dimensional grid sweep in order to minimize the error between computed equilibrium values and those obtained from the references. We started by scanning the parameter space for regions where the least squares error between data values and simulated values is minimal. We narrowed it down to the following: *α*_*t*_ ∈ [3, 4], *β*_*g*_ ∈ [0.0014, 0.0016] and *β*_*h*_ ∈ [0.0012, 0.0014]. The grid sweep was performed in MATLAB using ODE45. We also tested the model with other solvers (ODE23S, ODE15S) and obtained very similar results. Initial conditions (aside from *P* and *P*_*g*_) were randomly selected, since our focus was on equilibrium values.

We are left with calibrating *ρ* and *α*_*p*_. We started by replacing *S* = 0.71 by *S* + *P* = 0.71 at equilibrium. We have also replaced *I*_*g*_ + *I*_*gh*_ + *T*_*g*_ = 0.15 by *I*_*g*_ + *I*_*gh*_ + *T*_*g*_ + *P*_*g*_ = 0.15 and *I*_*g*_ = 0.1 by *I*_*g*_ + *P*_*g*_ = 0.1. Current estimates of PrEP users average to 5% [38, 39, 40, 41, 42]. Therefore, we added *P* + *P*_*g*_ = 0.05. When calibrating *ρ* and *α*_*p*_, we fixed the previously found baseline values for *α*_*t*_ = 3.875, *β*_*g*_ = 0.001475 and *β*_*h*_ = 0.001275. The baseline parameter values appear in Table 1.

## 3 Results

The simulation results are presented in the form of parameter planes and time series in the next two sub-sections.

### 3.1 Parameter planes

#### Increasing PrEP uptake rate increases gonorrhea prevalence

While PrEP prevents the transmission of HIV, it is ineffective against other STIs. Therefore, we examined the equilibrium prevalence of gonorrhea (*I*_*g*_ + *I*_*gh*_ + *P*_*g*_ + *T*_*g*_) for different rates of PrEP uptake. In particular, we were interested in optimal combinations of testing frequency for PrEP users (*f*_*p*_), and condom use or other preventive strategies (*α*_*p*_) in order to minimize the prevalence of gonorrhea.

We ran a two dimensional grid 101 × 101 of values for *f*_*p*_ ranging from 0.001 (testing roughly once every 3 years) to 0.04 (testing once every 25 days), and for *α*_*p*_ ranging from 0.0 (abstinence from sex, very high condom use or any other preventive strategy) to 20.0 (less precautions, lower condom use). We conducted this simulation for 4 different values of *ρ*: 0.0 (no PrEP uptake), 2.375 × 10^−6^ (baseline value), 3.562 × 10^−6^ (50% higher than baseline value) and 1.1875 × 10^−5^ (5 times higher than baseline value).

For the baseline parameter value of PrEP uptake (Fig 2a), we observe that an increase in risky behaviour (*α*_*p*_) causes a significant increase in gonorrhea prevalence. For example, the baseline increase in *α*_*p*_ from 1 to 12.75 causes an increase in gonorrhea prevalence from approximately 10 % to 16 %. On the other hand, an increase in the testing frequency (*f*_*p*_) causes a decrease in gonorrhea prevalence (for *α*_*p*_ unchanged). Lines of constant gonorrhea prevalence run approximately linearly across the parameter plane, such that the baseline increase in *α*_*p*_ would need to be accompanied by an increase in *f*_*p*_ far in excess of 0.04 (testing every 25 days). Unfortunately, this suggests that an increase in gonorrhea prevalence in a population where PrEP is widespread is very difficult to prevent, for realistic testing intervals.

**Figure 2:**
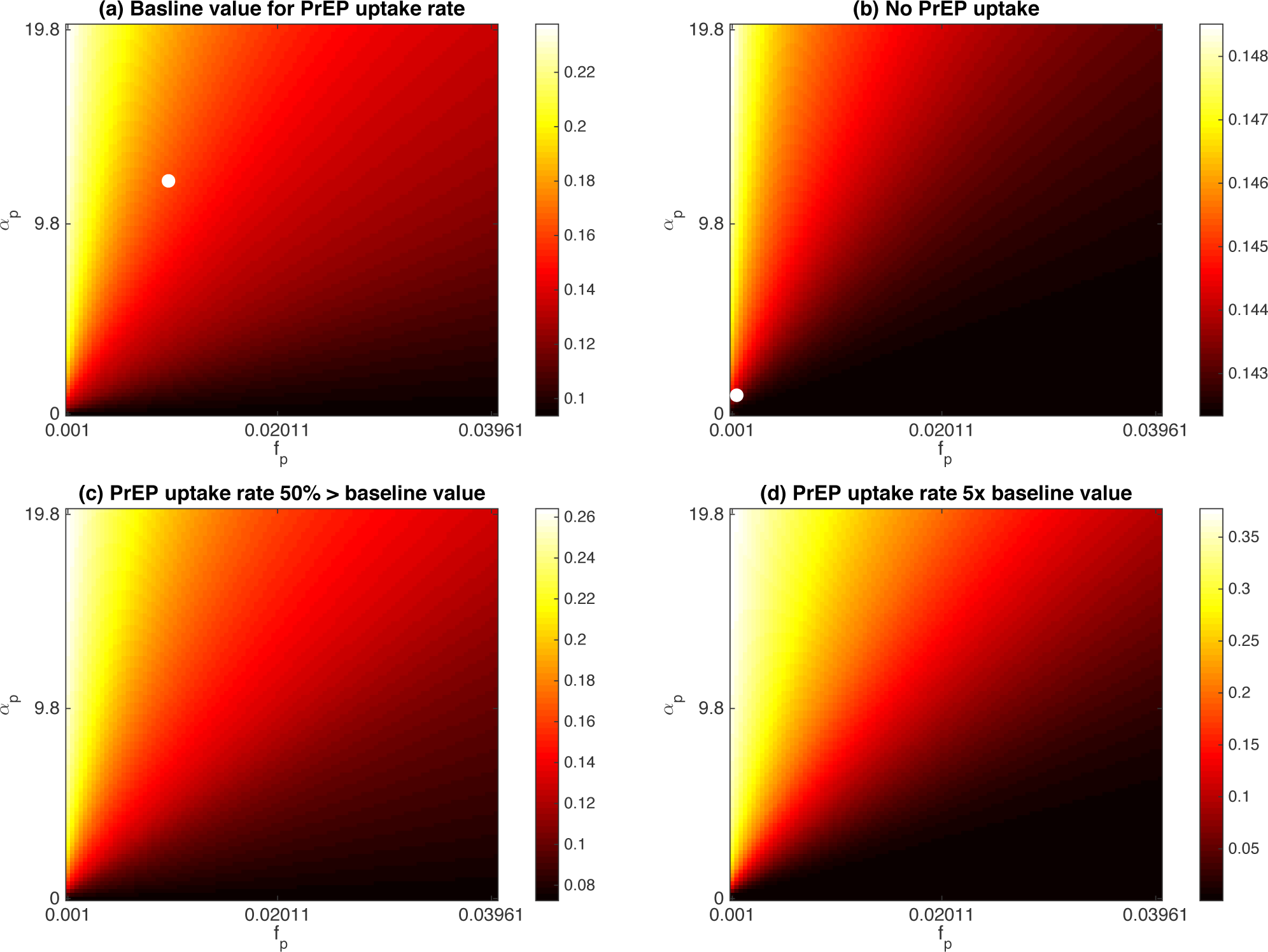
Gonorrhea prevalence heatmaps. Testing frequency for PrEP users (*f*_*p*_) and gonorrhea transmission risk factor for PrEP users (*α*_*p*_) are varied and all other parameters are held at baseline values. Different values of PrEP uptake rate (*ρ*) are considered. **(a)** PrEP uptake rate baseline value: *ρ* = 2.375 × 10^−6^. The white dot locates baseline values for *α*_*p*_ and *f*_*p*_. **(b)** No PreP uptake: *ρ* = 0.0. The white dot locates values for the pre-PrEP era: *f*_*p*_ = *f* and *α*_*p*_ = 1. **(c)** PrEP uptake rate 50% greater than baseline value: *ρ* = 3.562 × 10^−6^. **(d)** PrEP uptake rate 5 times greater than baseline value: *ρ* = 1.1875 × 10^−5^. Black regions represent regions of lower gonorrhea prevalence. Yellow regions represent regions of higher gonorrhea prevalence. Note the difference in vertical scale between the four subpanels.

When *ρ* = 0 (no PrEP uptake, Fig 2b), we observe a slight variation of about 0.5% in the prevalence of gonorrhea in the (*f*_*p*_, *α*_*p*_) plane, as expected (note the vertical scale of subpanel 2b). This is due to the fact that we have chosen non-zero initial conditions for *P* and *P*_*g*_. In particular, we have set *P* (0) = 0.08 and *P*_*g*_(0) = 0.02. We have examined the case when *P* (0) = *P*_*g*_(0) = *ρ* = 0 and there was no variation in the levels of gonorrhea in the population.

With information propagating through social media and awareness campaigns, more individuals are becoming aware of PrEP and its efficacy in HIV prevention. We suspect that the PrEP uptake rate will be on the rise within the next decade. We have therefore investigated scenarios where the uptake rate is increased by 50% from the baseline value (Fig 2c) and is 5 times higher than the baseline value (Fig 2d). This inevitable increase in *ρ* can be potentially problematic since greater increases in the prevalence of gonorrhea are observed for higher values of *ρ*. For instance, when *ρ* is 5 times larger than the baseline value, gonorrhea levels can reach 35% of the entire population if no precautions are carefully taken into account (yellow regions in Fig 2c,d).

On the other hand, this increase in the uptake rate offers a wider area in the (*f*_*p*_, *α*_*p*_) parameter plane where gonorrhea prevalence remains low (compare black regions in Fig 2c and Fig 2d). In fact, under these scenarios of higher PrEP uptake, the testing frequency does not need to be increased as much as under the baseline PrEP uptake, in order for gonorrhea prevalence to remain unchanged. This flexibility allows for better control and the prevention of outbreaks. If *α*_*p*_ is too high, the focus would be on increasing the testing frequency to maintain sub-epidemic levels of gonorrhea in the population.

We also noticed a linear border between regions of higher and lower gonorrhea prevalence. The border separating yellow regions from red regions is much steeper than that separating red regions from black regions. This is an indication of the importance in maintaining a minimum testing frequency to keep gonorrhea from infecting a greater proportion of the population.

#### Sufficiently frequent STI testing controls gonorrhea prevalence

Before starting a PrEP regimen, every individual needs to go through a series of tests. The individual should test negative for HIV as a first step to qualify. They also have to test for common STIs such as gonorrhea, chlamydia, and syphilis. If the results come back positive for any of the STIs, an anti-bacterial treatment is prescribed first, and pending recovery, PrEP qualification is revisited. Finally, testing the liver and kidney functions is also essential before starting PrEP. The pills can have damaging effects on the liver [43]. If ALT (alanine aminotransaminase) and AST (aspartate transaminase) levels are too high, a nurse practitioner (or a physician) recommends lowering the levels before starting PrEP.

Public health agencies in Canada have set testing frequency for PrEP users to be once every 3 months [44]. This is standard follow up procedure and PrEP is only prescribed for 3 months only with no refills. A PrEP user needs to visit their physician and test negative for all STIs before they receive another 3 months prescription. Testing is usually subsidized by local governments or paid for by insurance companies (in the case of very specific tests). In this section, we examine several scenarios of testing frequency ranging from once every 2 months (more frequent than the current recommended frequency), twice per year, and once every 5 years. We run a 2-dimensional 101 × 101 grid sweep of the two parameters *ρ* and *α*_*p*_. The parameter *ρ* ranges from 10^−6^ to 5 × 10^−5^. The parameter *α*_*p*_ ranges from 0 to 20.

The parameter planes illustrate that frequent testing is essential in order to maintain a lower prevalence of gonorrhea. Gonorrhea infections can reach levels as high as 60% of the population when individuals on PrEP are tested only once every 5 years (Fig 3d) and 50% when tested twice every year (Fig 3b). Under both testing scenarios, black regions are very thin and therefore we would require extreme measures to control the propagation of the disease.

**Figure 3:**
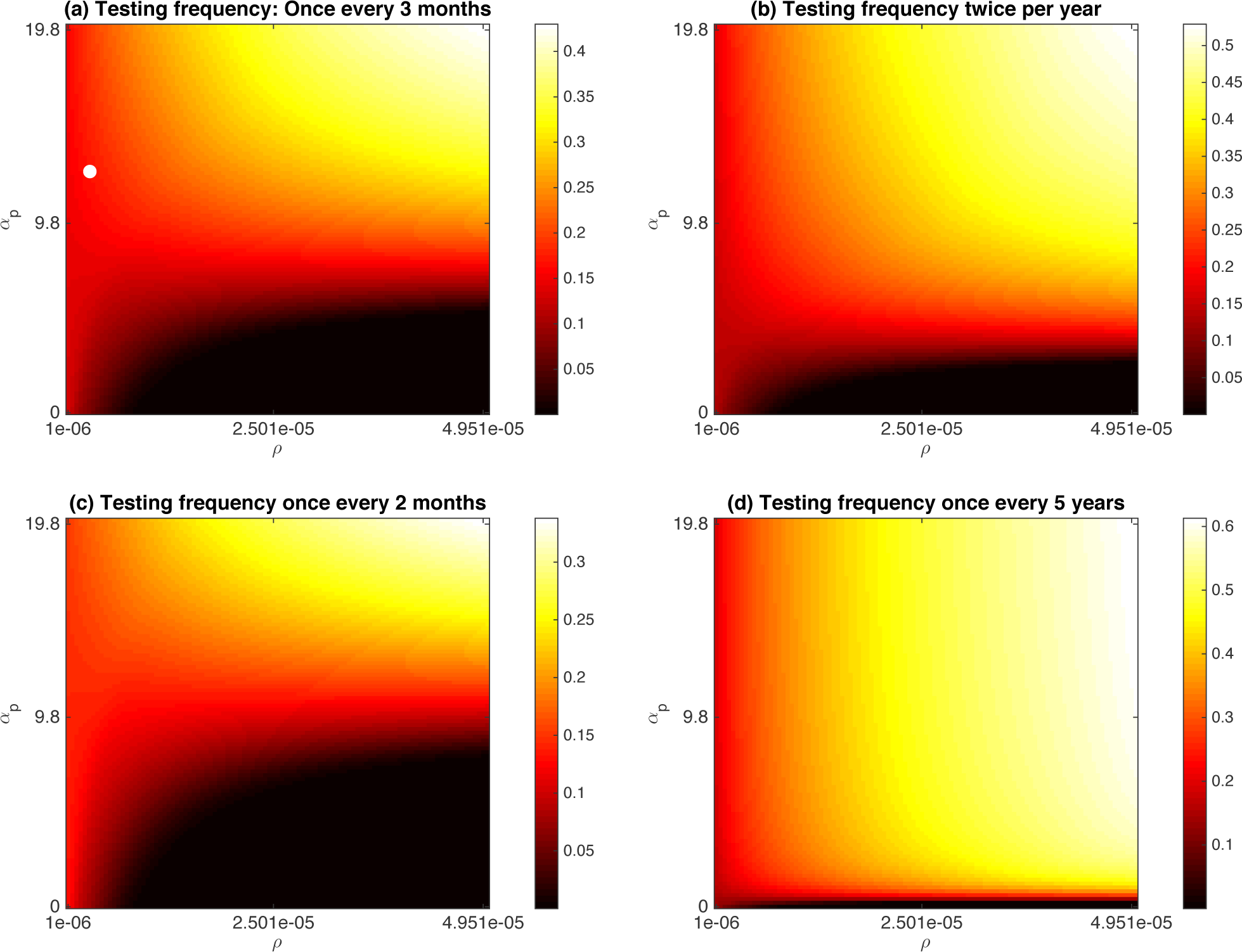
Gonorrhea prevalence heatmaps. PrEP uptake rate (*ρ*) and gonorrhea transmission risk factor for PrEP users (*α*_*p*_) are varied and all other parameters are held at baseline values. Different values of testing frequency for PrEP users (*f*_*p*_) are considered. **(a)** Testing frequency once every 3 months (baseline value): *f*_*p*_ = 1*/*90 ≈ 0.0111. The white dot locates baseline values for *α*_*p*_ and *ρ*. **(b)** Testing frequency twice every year: *f*_*p*_ = 1*/*182 ≈ 0.0055. **(c)** Testing frequency once every two months: *f*_*p*_ = 1*/*60 ≈ 0.0167. **(d)** Testing frequency once every 5 years: *f*_*p*_ = 1*/*1825 ≈ 0.0005. Black regions represent regions of lower gonorrhoea prevalence. Yellow regions represent regions of higher gonorrhea prevalence. Note the difference in vertical scale between the four subpanels.

On the other hand, testing once every 2 months (Fig 3c) not only minimizes gonorrhea infections but allows for more flexibility in parameter combinations to maintain low gonorrhea prevalence. The black region in Fig 3c indicates that despite lower than necessary levels of condom use, we can still keep gonorrhea in control and prevent outbreaks at baseline PrEP uptake, as long as risky behaviour does not increase too much under PrEP (*α*_*p*_;S 8). However, more frequent testing may be cost prohibitive for ministries of health. Also, more frequent testing may reduce adherence since PrEP users would be required to visit their physician more often and do more blood work, in this case 6 times per year (versus 4 times per year under current recommendation).

The barrier between darker regions and lighter regions is nonlinear in these parameter planes. Moreover, the parameter planes show a region of rapid increase in gonorrhea prevalence as risky behaviour (*α*_*p*_) increases from the pre-PrEP value to the baseline PrEP value of 12.8, for realistic testing frequencies. This shows that small increases in sexually risky behaviour may not change gonorrhea prevalence very much although a further increase could shift the system into a region where gonorrhea prevalence increases suddenly. Hence we expect that increases in gonorrhea prevalence under PrEP may be highly dependent on the population under consideration. Also, this emphasizes the importance of increasing the usage of preventive strategies such as condoms.

#### Increasing condom usage in combination with increasing the testing frequency is the most effective and realistic way to control gonorrhoea

Here we constructed a 101 × 101 grid for *ρ* ranging from 10^−6^ to 5 × 10^−5^ and *f*_*p*_ ranging from 0.001 to 0.04 under four different scenarios for the value of *α*_*p*_, representing the increase in risky behaviour after introduction of PrEP due to decreased condom usage, for instance. Recall that *α*_*p*_ = 1 corresponds to no change in risky behaviour due to PrEP, while *α*_*p*_ *ϵ;* 1 represents an increase in risky behaviour. *α*_*p*_ = 6.375 represents an increase in condom use (or other preventive strategies) by 50% compared to the baseline value *α*_*P*_ = 12.8, whereas *α*_*p*_ = 25.5 represents a 50% decrease. *α*_*p*_ = 63.75 represents a 5 times decrease in condom use.

These parameter planes confirm some of the observations of Figure 3. For instance, gonorrhea prevalence decreases sharply for an intermediate range of values for the testing frequency *f*_*p*_. Most importantly, the parameter planes show that a combination of decreasing risky behaviour through greater condom usage relatively to the baseline scenario, and changing testing frequency to once every two months (Figure 4b), can keep gonorrhea prevalence close to pre-PrEP levels. This is crucial since, as pointed out in the previous subsection, high frequency testing may be cost prohibitive and/or may incur low adherence rates. An interesting feature of Figure 4b is the switch of location of black regions from lower *ρ* and higher *f*_*p*_ values when *α*_*p*_ is greater than the baseline value to higher *ρ* and lower *f*_*p*_ values when *α*_*p*_ is lower than the baseline value (Figure 4a,b versus Figure 4c,d). This furthermore indicates that condom use can be very critical in determining optimal policies to reduce gonorhhea prevalence.

**Figure 4:**
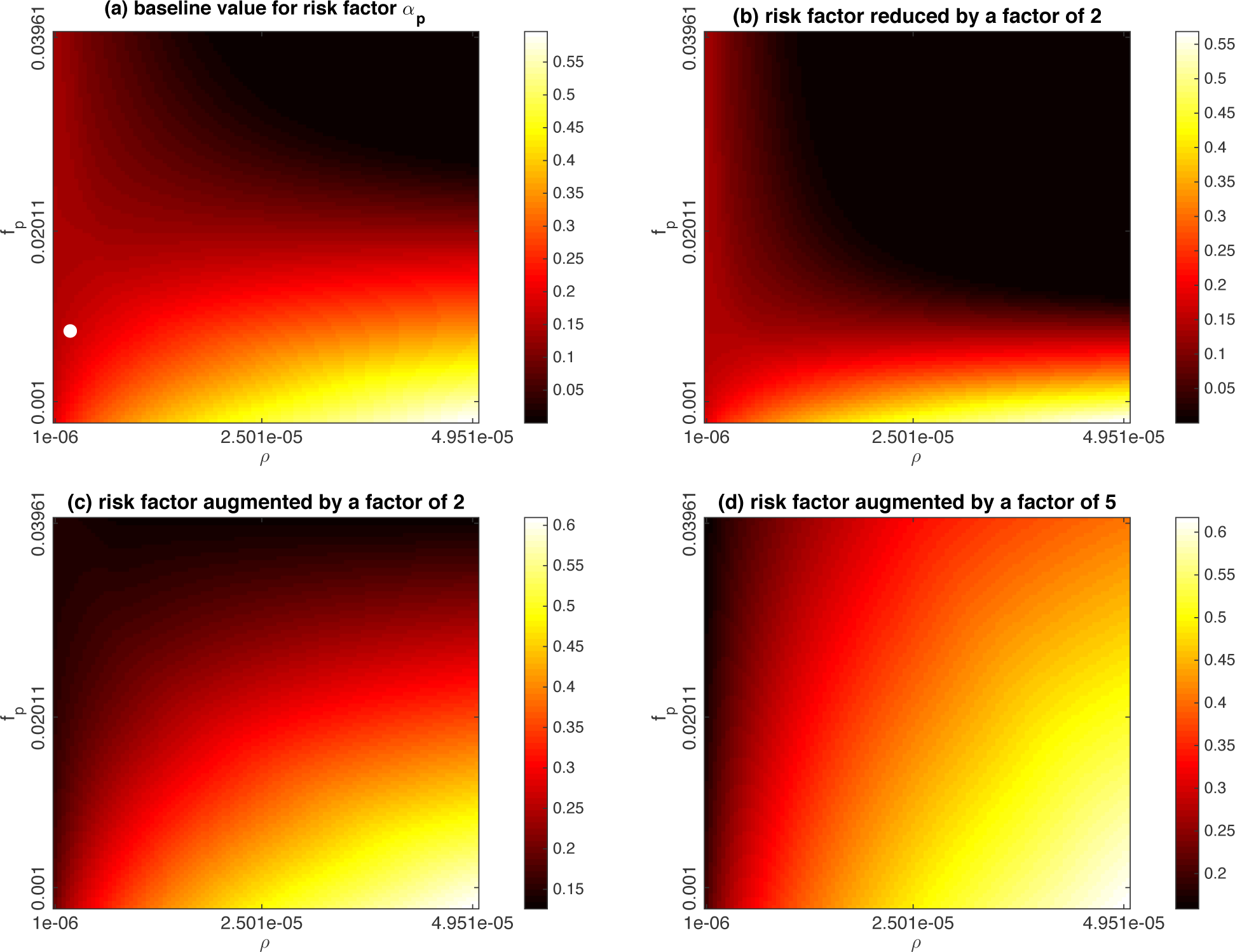
Gonorrhea prevalence heatmaps. PrEP uptake rate (*ρ*) and testing frequency for PrEP users (*f*_*p*_) are varied and all other parameters are held at baseline values. Different values of gonorrhea transmission risk factor for PrEP users (*α*_*p*_) are considered. **(a)** gonorrhea transmission risk factor for PrEP users baseline value: *α*_*p*_ = 12.75. The white dot locates baseline values for *f*_*p*_ and *ρ*. **(b)** gonorrhea transmission risk factor for PrEP users reduced by a factor of 2: *α*_*p*_ = 6.375. **(c)** gonorrhea transmission risk factor for PrEP users augmented by a factor of 2: *α*_*p*_ = 25.5. **(d)** gonorrhea transmission risk factor for PrEP users multiplied by 5: *α*_*p*_ = 63.75. Black regions represent regions of lower gonorrhea prevalence. Yellow regions represent regions of higher gonorrhea prevalence. Note the difference in vertical scale between the four subpanels.

Contrariwise, very low rates of condom usage (high *α*_*p*_, Fig 4c,d) result in very high prevalence of gonorrhea, unless the testing frequency is impractically high. Hence, condom use remains essential in the prevention of other transmitted STIs such as gonorrhea, despite increased STI testing frequency when individuals start PrEP.

### 3.2 Time series analysis

Time series indicate the temporal evolution of HIV and GC prevalence, showing that changes in prevalence unfold on different time scales depending on both the diseaes and the intervention scenarios (Figs 5, 6). Upon the introduction of PrEP, HIV prevalence decreases exponentially from 4% to lower than 0.1% within 50 years, consistent with experience on the ground with PrEP programs (Fig 6a).

**Figure 5:**
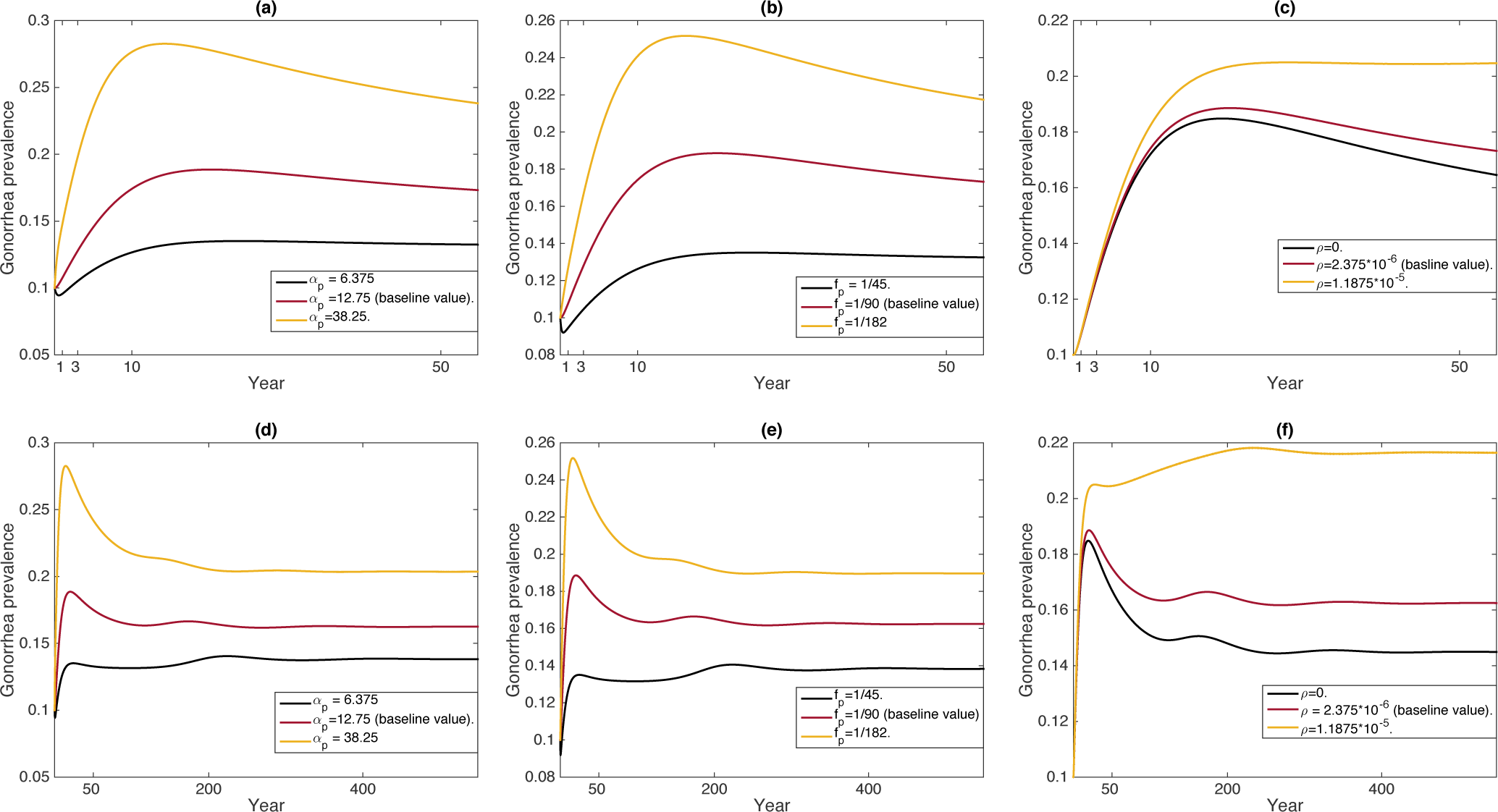
Time series of gonorrhea prevalence over different time scales. In each subfigure, all parameters but one are kept at baseline values. **(a**,**d)** gonorrhea transmission risk factor for PrEP users (*α*_*p*_) is varied: baseline value in red (12.75), reduced by a factor of 2 in black (6.375), augmented by a factor of 3 in yellow (38.25). **(b**,**e)** Testing frequency for PrEP users (*f*_*p*_) is varied: baseline value in red (1*/*90: once every 3 months), testing once every 1.5 months in black (1*/*45), testing twice per year in yellow (1*/*182). **(c**,**f)** PrEP uptake rate (*ρ*) is varied: baseline value in red (2.375 × 10^−6^), No PrEP uptake in black (0), PrEP uptake rate 5 times greater than baseline value in yellow (1.1875 × 10^−5^).

**Figure 6:**
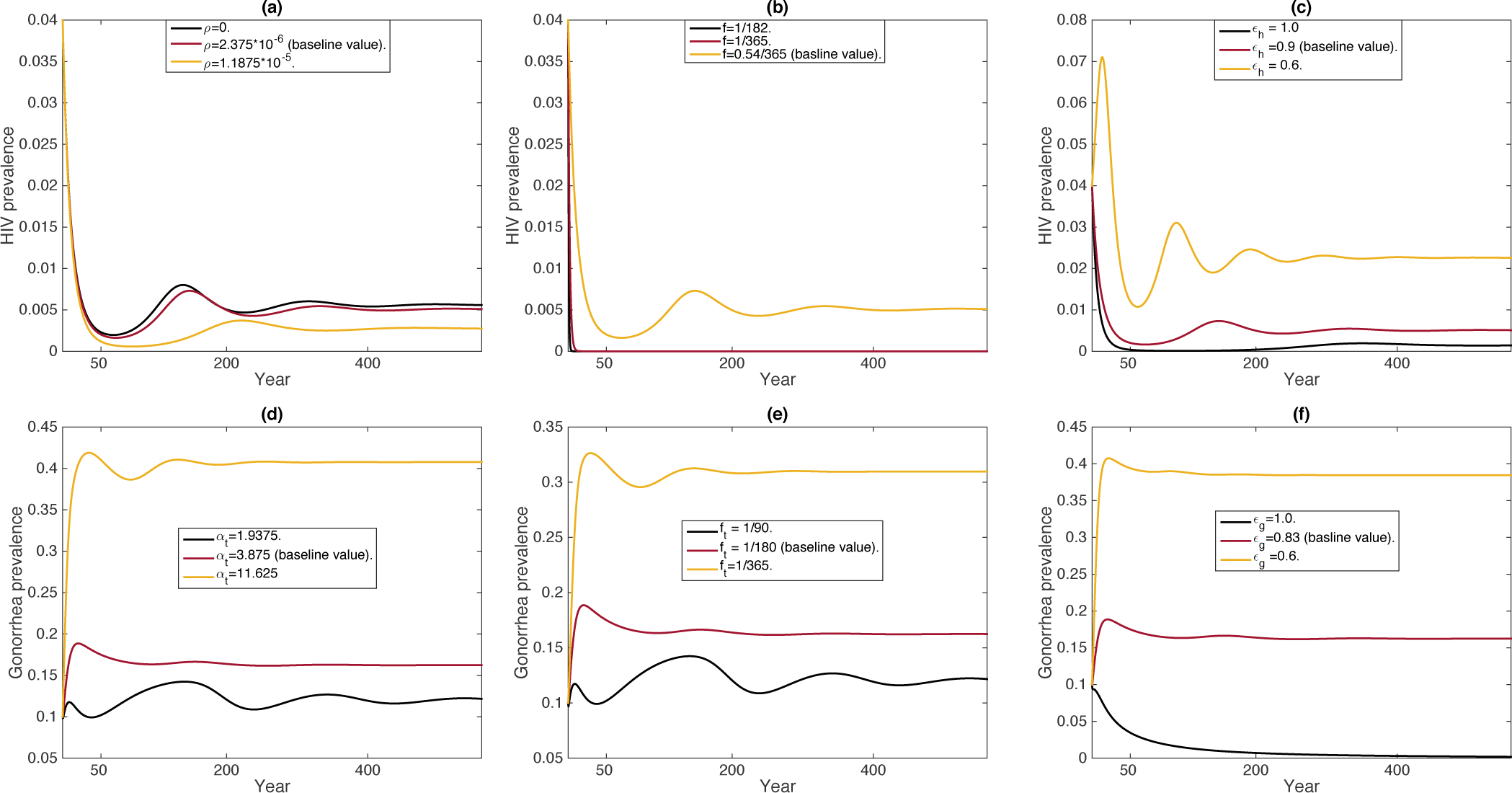
Time series of gonorrhea and HIV prevalence over long time scales. In the first row, we compute HIV prevalence. We vary **(a)** PrEP uptake rate *ρ* (no PrEP uptake *ρ* = 0, basline value PrEP uptake rate and 5 times baseline value), **(b)** testing frequency for individuals in the susceptible class *f* (once every two years, once every year, twice per year), **(c)** efficacy of HIV treatment *ϵ*_*h*_ (90% effective:baseline value, 100% effective, 60% effective). In the second row, we compute gonorrhea prevalence. We vary **(d)** gonorrhea transmission risk factor for individuals who are treated for HIV (*α*_*t*_ = 3.875 baseline value, twice lower than baseline value, three times higher than baseline value), **(e)** testing frequency for individuals who are treated for HIV *f*_*t*_ (twice per year, once per year, once every 3 months), **(f)** efficacy of gonorrhea treatment *ϵ*_*g*_ (83% effective: baseline value, 100% effective, 60% effective).

Exponential behaviour, as opposed to oscillatory behaviour, is also observed for gonorrhea. All simulations have resulted in a spike in gonorrhea followed by an exponential decrease to stable values (Fig 5,6). The values depend on the choice of social parameters. There are exceptions to this trend. When PrEP uptake rate is five times greater than baseline value, gonorrhea prevalence increases steadily until it reaches a stable value of about 22% (Fig 5f). The baseline value for PrEP uptake rate reflects the current trends in PrEP awareness. PrEP awareness is increasing with information being dissipated through social networks. Immediate decline in gonorrhea prevalence, despite current PrEP uptake rate, is obtained when the efficacy of gonorrhea treatment is 100% (Fig 6f). The efficacy of treatment is related to early diagnosis and other biological factors. Interestingly, there is a situation under which we observe oscillatory behaviour in gonorrhea prevalence. If condom use is doubled for individuals who are treated for HIV, or if they get tested twice as often, gonorrhea prevalence oscillates (Fig 6d,e black curves).

Gonorrhea prevalence increases to a maximum within a decade and then it starts dropping to a stable value (Fig 5,6). The maximum increase depends on the combination of parameters. Highest levels of gonorrhea prevalence range between 20% and 25%. This represents an increase of up to 60% from current values. Those levels are observed if people on PrEP get tested less frequently, for example twice per year (Fig 5b,e) or if their condom use is reduced by a factor of 3 (Fig 5a,d). The interesting observation, however, is that the period of time until gonorrhea prevalence decreases is uniform for many parameter combinations. Even if people on PrEP get tested once every month and a half or they start using condoms more often, we still observe an initial spike in gonorrhea prevalence (black curves in Fig 5a,b).

There exist a value of PrEP uptake rate somewhere between the baseline value and 5 times the baseline value where the qualitative behaviour of gonorrhea prevalence changes (Fig 5c,f). This indicates, that under current social behaviour, if PrEP uptake increases, then there would be a 70% increase in gonorrhea prevalence (from 16% to 22%) at equilibrium value.

Testing frequency is essential to maintain lower levels of gonorrhea prevalence. Individuals on PrEP, as well as those who are treated for HIV, must get tested frequently otherwise gonorrhea prevalence rises (Fig 6e). While there are no testing policies and requirements for individuals treated for HIV, simulations show that when they get tested once every 3 months versus once per year, gonorrhea prevalence falls by 60%. This shows that gonorrhea prevalence is not only affected by individuals on PrEP but by other members of the community as well. The average testing frequency for gay and bisexual men is roughly once every two years in some populations, which is considered very low. PrEP is not the only way to prevent the transmission of HIV. Even though HIV is experiencing a decline [45, 46, 47], if members of the community are tested on average once per year (Fig 6b red curve), HIV will decline further.

## 4 Discussion

Here we developed and analyzed a compartmental model of HIV and gonorrhea transmission including allowances for behavioural changes upon the introduction of pre-exposure prophylaxis (PrEP) against HIV infection. We found that gonorrhea prevalence increases for a wide range of parameter assumptions, due to a likely increase in risky behaviour from the introduction of PrEP. Our results show that STI testing for individuals on PrEP once every 3 months is not enough to prevent an increase in gonorrhea prevalence, for our baseline assumptions about the impact of PrEP on risky behaviour. Moreover, increased STI testing frequency alone is not enough to prevent a rise in gonorrhea prevalence because the testing frequency would have to be increased to impractical levels in order to be effective. However, testing somewhat more frequently than every 3 months, in combination with sufficient condom usage by individuals on PrEP, would be successful in maintaining gonorrhea prevalence at pre-PrEP levels.

Our model highlighted the importance of testing freqency. Testing frequency in many populations without mechanisms for recall (e.g. postal or phone reminders about when it is time to get tested) can be as low as once every two years. Our model predictions show that massive reductions in prevalence are possible when moving from testing every two years to testing every year. This suggests it might be worthwhile to better incentivize frequency STI testing.

Our findings about the importance of both condoms and STI testing to keep gonorrhea under control in the PrEP era do not replace other good advice. For instance, STI testing once every 6 months is recommended for individuals being treated for HIV infection every time they have a follow up with their physician on their viral loads and overall health. Other preventive strategies include having regular partners, asking sexual partners about their testing frequency and sexual habits, abstinence, adhering to treatment protocol when infected by an STI, and informing sexual partners when infected with an STI.

We developed this model for gonorrhea, although the model assumptions are similar to those that would be made for other bacterial STIs such as chlamydia or syphilis. Hence, we expect qualitatively similar findings for all bacterial STIs.

Our model made simplifying assumptions that could influence its predictions. For instance, gonorrhea and HIV, like many pathogens, are currently developing more drug resistance [48, 49, 50, 51, 52]. Gonorrhea treatment efficacy is connected to early diagnosis and adherence to the medication [53]. Hence if treatment for gonorrhea becomes less effective due to drug resistance, gonorrhea prevalence in many populations will rise more than our model has predicted.

Similarly, both social dynamics and contact network structure can play a significant role in the spread of infectious diseases, including HIV and gonorrhea [27, 28, 54, 54, 55, 56, 57, 58, 59, 60, 61]. In future work, our study design could be refined by using network simulations that account for social dynamics concerning HIV and gonorrhea transmission. The continued persistence of HIV in the gay and bisexual men’s community, along with the resurgence of gonorrhea and other bacterial STIs, suggests a continued and urgent need for mathematical modelling studies that can help inform recommendations to result in better infection control.

## Data Availability

The code used to generate the simulation results is available upon request to the authors.

## References

[1] Miriam Smith. Social movements and equality seeking: the case of gay liberation in canada. Canadian Journal of Political Science/Revue canadienne de science politique, 31(2):285–309, 1998.

[2] Allan Bérubé. The history of gay bathhouses. Journal of Homosexuality, 44(3-4):33–53, 2003.

[3] Feng Gao, Elizabeth Bailes, David L Robertson, Yalu Chen, Cynthia M Rodenburg, Scott F Michael, Larry B Cummins, Larry O Arthur, Martine Peeters, George M Shaw, et al. Origin of hiv-1 in the chimpanzee pan troglodytes troglodytes. Nature, 397(6718):436, 1999.

[4] Andrew Rambaut, David L Robertson, Oliver G Pybus, Martine Peeters, and Edward C Holmes. Phylogeny and the origin of hiv-1. Nature, 410(6832):1047, 2001.

[5] Chris Beyrer, Stefan D Baral, Frits Van Griensven, Steven M Goodreau, Suwat Chariyalertsak, Andrea L Wirtz, and Ron Brookmeyer. Global epidemiology of hiv infection in men who have sex with men. the Lancet, 380(9839):367–377, 2012.

[6] Louis M Mansky. Retrovirus mutation rates and their role in genetic variation. Journal of general Virology, 79(6):1337–1345, 1998.

[7] Rachel A Royce, Arlene Sena, Willard Cates Jr, and Myron S Cohen. Sexual transmission of hiv. New England Journal of Medicine, 336(15):1072–1078, 1997.

[8] Steven D Pinkerton and Paul R Abramson. Effectiveness of condoms in preventing hiv transmission. Social science & medicine, 44(9):1303–1312, 1997.

[9] Susan C Weller. A meta-analysis of condom effectiveness in reducing sexually transmitted hiv. Social science & medicine, 36(12):1635–1644, 1993.

[10] W Cates Jr and H Hunter Handsfield. Hiv counseling and testing: does it work? American Journal of Public Health, 78(12):1533–1534, 1988.

[11] Ashley York. Undetectable equals untransmittable, 2019.

[12] Kimberly A Workowski and Gail A Bolan. Sexually transmitted diseases treatment guidelines, 2015. MMWR. Recommendations and reports: Morbidity and mortality weekly report. Recommendations and reports, 64(RR-03):1, 2015.

[13] John Tapsall. Antibiotic resistance in neisseria gonorrhoeae is diminishing available treatment options for gonorrhea: some possible remedies. Expert review of anti-infective therapy, 4(4):619–628, 2006.

[14] Seth C Kalichman, Jennifer Pellowski, and Christina Turner. Prevalence of sexually transmitted coinfections in people living with hiv/aids: systematic review with implications for using hiv treatments for prevention. Sexually transmitted infections, 87(3):183–190, 2011.

[15] Lynn A Paxton, Tony Hope, and Harold W Jaffe. Pre-exposure prophylaxis for hiv infection: what if it works? The lancet, 370(9581):89–93, 2007.

[16] Christopher B Hurt, Joseph J Eron Jr, and Myron S Cohen. Pre-exposure prophylaxis and antiretroviral resistance: Hiv prevention at a cost? Clinical Infectious Diseases, 53(12):1265–1270, 2011.

[17] Sheena McCormack, David T Dunn, Monica Desai, David I Dolling, Mitzy Gafos, Richard Gilson, Ann K Sullivan, Amanda Clarke, Iain Reeves, Gabriel Schembri, et al. Pre-exposure prophylaxis to prevent the acquisition of hiv-1 infection (proud): effectiveness results from the pilot phase of a pragmatic open-label randomised trial. The Lancet, 387(10013):53–60, 2016.

[18] Anna Van Laarhoven, Elske Hoornenborg, Roel Achterbergh, Henry De Vries, Maria Prins, and Maarten Schim Van Der Loeff. O09. 3 changes in sexual risk behaviour among daily prep users after 6 months of use in the amsterdam prep project, 2017.

[19] Kamair Alaei, Christopher A Paynter, Shao-Chiu Juan, and Arash Alaei. Using preexposure prophylaxis, losing condoms? preexposure prophylaxis promotion may undermine safe sex. Aids, 30(18):2753–2756, 2016.

[20] Florence M Koechlin, Virginia A Fonner, Sarah L Dalglish, Kevin R O’Reilly, Rachel Baggaley, Robert M Grant, Michelle Rodolph, Ioannis Hodges-Mameletzis, and Caitlin E Kennedy. Values and preferences on the use of oral pre-exposure prophylaxis (prep) for hiv prevention among multiple populations: a systematic review of the literature. AIDS and Behavior, 21(5):1325–1335, 2017.

[21] Michael W Traeger, Sophia E Schroeder, Edwina J Wright, Margaret E Hellard, Vincent J Cornelisse, Joseph S Doyle, and Mark A Stoové. Effects of pre-exposure prophylaxis for the prevention of human immunodeficiency virus infection on sexual risk behavior in men who have sex with men: a systematic review and meta-analysis. Clinical Infectious Diseases, 67(5):676–686, 2018.

[22] M Hull and DHS Tan. Can we eliminate hiv?: Setting the stage for expanding hiv pre-exposure prophylaxis use in canada. Canada Communicable Disease Report, 43(12):272, 2017.

[23] Oluwaseun Sharomi, C Podder, Abba Gumel, and Baojun Song. Mathematical analysis of the transmission dynamics of hiv/tb coinfection in the presence of treatment. Mathematical Biosciences and Engineering, 5(1):145, 2008.

[24] El Mehdi Lotfi, Marouane Mahrouf, Mehdi Maziane, Cristiana J Silva, Delfim FM Torres, and Noura Yousfi. A minimal hiv-aids infection model with general incidence rate and application to morocco data. arXiv preprint 1812.06965, 2018.

[25] Narat Punyacharoensin, William John Edmunds, Daniela De Angelis, Valerie Delpech, Graham Hart, Jonathan Elford, Alison Brown, Noel Gill, and Richard G White. Modelling the hiv epidemic among msm in the united kingdom: quantifying the contributions to hiv transmission to better inform prevention initiatives. Aids, 29(3):339–349, 2015.

[26] Cristiana J Silva and Delfim FM Torres. A sica compartmental model in epidemiology with application to hiv/aids in cape verde. Ecological complexity, 30:70–75, 2017.

[27] Stephen Tully, Monica-Gabriela Cojocaru, and Chris T Bauch. Multiplayer games and hiv transmission via casual encounters. Mathematical Biosciences & Engineering, 14(2):359–376, 2017.

[28] Stephen Tully, Monica Cojocaru, and Chris T Bauch. Sexual behavior, risk perception, and hiv transmission can respond to hiv antiviral drugs and vaccines through multiple pathways. Scientific reports, 5:15411, 2015.

[29] Brian G Williams, James O Lloyd-Smith, Eleanor Gouws, Catherine Hankins, Wayne M Getz, John Hargrove, Isabelle De Zoysa, Christopher Dye, and Bertran Auvert. The potential impact of male circumcision on hiv in sub-saharan africa. PLoS medicine, 3(7):e262, 2006.

[30] Robert J Smith and Sally M Blower. Could disease-modifying hiv vaccines cause population-level perversity? The Lancet infectious diseases, 4(10):636–639, 2004.

[31] Linda Gao and Herbert Hethcote. Simulations of rubella vaccination strategies in china. Mathematical biosciences, 202(2):371–385, 2006.

[32] Christoph D Spinner, Christoph Boesecke, Alexander Zink, Heiko Jessen, Hans-Jürgen Stellbrink, Jürgen Kurt Rockstroh, and Stefan Esser. Hiv pre-exposure prophylaxis (prep): a review of current knowledge of oral systemic hiv prep in humans. Infection, 44(2):151–158, 2016.

[33] Alison J Rodger, Valentina Cambiano, Tina Bruun, Pietro Vernazza, Simon Collins, Olaf Degen, Giulio Maria Corbelli, Vicente Estrada, Anna Maria Geretti, Apostolos Beloukas, et al. Risk of hiv transmission through condomless sex in serodifferent gay couples with the hiv-positive partner taking suppressive antiretroviral therapy (partner): final results of a multicentre, prospective, observational study. The Lancet, 2019.

[34] Jalal Poorolajal, Leila Molaeipoor, Minoo Mohraz, Hossein Mahjub, Maryam Taghizadeh Ardekani, Pegah Mirzapour, and Hanieh Golchehregan. Predictors of progression to aids and mortality post-hiv infection: a long-term retrospective cohort study. AIDS care, 27(10):1205–1212, 2015.

[35] Centre for Communicable Diseases, Infectious Disease Prevention Infection Control, and Public Health Agency of Canada Control Branch. Public health agency of canada. m-track: Enhanced surveillance of hiv, sexually transmitted and blood-borne infections, and associated risk behaviours among men who have sex with men in canada. phase 1 report. 2011.

[36] Banks P. Marchand R. Robert W. Gustafson R. Hogg R. Gilbert M. Trussler, T. and the ManCount Survey Team. Mancount sizes-up the gaps: a sexual health survey of gay men in vancouver. vancouver coastal health: Vancouver. 2010.

[37] Charlotte K Kent, Janice K Chaw, William Wong, Sally Liska, Steven Gibson, Gregory Hubbard, and Jeffrey D Klausner. Prevalence of rectal, urethral, and pharyngeal chlamydia and gonorrhea detected in 2 clinical settings among men who have sex with men: San francisco, california, 2003. Clinical Infectious Diseases, 41(1):67–74, 2005.

[38] Jayoti Rana, James Wilton, Shawn Fowler, Trevor A Hart, Ahmed M Bayoumi, and Darrell HS Tan. Trends in the awareness, acceptability, and usage of hiv pre-exposure prophylaxis among at-risk men who have sex with men in toronto. Canadian Journal of Public Health, pages 1–11, 2018.

[39] Terrance Mosley, Moliehi Khaketla, Heather L Armstrong, Zishan Cui, Paul Sereda, Nathan J Lachowsky, Mark W Hull, Gbolahan Olarewaju, Jody Jollimore, Joshua Edward, et al. Trends in awareness and use of hiv prep among gay, bisexual, and other men who have sex with men in vancouver, canada 2012–2016. AIDS and Behavior, pages 1–16, 2018.

[40] Phillip L Hammack, Ilan H Meyer, Evan A Krueger, Marguerita Lightfoot, and David M Frost. Hiv testing and pre-exposure prophylaxis (prep) use, familiarity, and attitudes among gay and bisexual men in the united states: A national probability sample of three birth cohorts. PloS one, 13(9):e0202806, 2018.

[41] William C Goedel, Kenneth H Mayer, Matthew J Mimiaga, and Dustin T Duncan. Considerable interest in pre-exposure prophylaxis uptake among men who have sex with men recruited from a popular geosocial-networking smartphone application in london. Global public health, pages 1–10, 2017.

[42] Rudy Patrick, David Forrest, Gabriel Cardenas, Jenevieve Opoku, Manya Magnus, II Gregory Phillips, Alan Greenberg, Lisa Metsch, Michael Kharfen, Marlene LaLota, et al. Awareness, willingness, and use of pre-exposure prophylaxis among men who have sex with men in washington, dc and miami-dade county, fl: national hiv behavioral surveillance, 2011 and 2014. JAIDS Journal of Acquired Immune Deficiency Syndromes, 75:S375–S382, 2017.

[43] Raymond A Tetteh, Barbara A Yankey, Edmund T Nartey, Margaret Lartey, Hubert GM Leufkens, and Alexander NO Dodoo. Pre-exposure prophylaxis for hiv prevention: safety concerns. Drug safety, 40(4):273–283, 2017.

[44] Darrell HS Tan, Mark W Hull, Deborah Yoong, Cécile Tremblay, Patrick O’byrne, Réjean Thomas, Julie Kille, Jean-Guy Baril, Joseph Cox, Pierre Giguere, et al. Canadian guideline on hiv pre-exposure prophylaxis and nonoccupational postexposure prophylaxis. Canadian Medical Association Journal, 189(47):E1448–E1458, 2017.

[45] Nneka Nwokolo, Andrew Hill, Alan McOwan, and Anton Pozniak. Rapidly declining hiv infection in msm in central london. The lancet HIV, 4(11):e482–e483, 2017.

[46] Eric PF Chow, Nicholas A Medland, Ian Denham, Edwina J Wright, and Christopher K Fairley. Decline in new hiv diagnoses among msm in melbourne. The Lancet HIV, 5(9):e479–e481, 2018.

[47] K Tomas, P Dhami, C Houston, S Ogunnaike-Cooke, and C Rank. Good news on hiv: Hiv in canada: 2009 to 2014. Canada Communicable Disease Report, 41(12):292, 2015.

[48] Emilie Alirol, Teodora E Wi, Manju Bala, Maria Luiza Bazzo, Xiang-Sheng Chen, Carolyn Deal, Jo-Anne R Dillon, Ranmini Kularatne, Jutta Heim, Rob Hooft van Huijsduijnen, et al. Multidrugresistant gonorrhea: A research and development roadmap to discover new medicines. PLoS medicine, 14(7):e1002366, 2017.

[49] I Martin, P Sawatzky, V Allen, B Lefebvre, LMN Hoang, P Naidu, J Minion, P Van Caeseele, D Haldane, RR Gad, et al. Multidrug-resistant and extensively drug-resistant gonorrhea in canada, 2012–2016. 2019.

[50] Tomas Cihlar and Marshall Fordyce. Current status and prospects of hiv treatment. Current opinion in virology, 18:50–56, 2016.

[51] Emma Cunningham, Yuen-Ting Chan, Adamma Aghaizu, David F Bibby, Gary Murphy, Jennifer Toss- will, Ross J Harris, Richard Myers, Nigel Field, Valerie Delpech, et al. Enhanced surveillance of hiv-1 drug resistance in recently infected msm in the uk. Journal of Antimicrobial Chemotherapy, page dkw404, 2016.

[52] Genevieve Rocheleau, Conrado Franco-Villalobos, Natalia Oliveira, Zabrina L Brumme, Melanie Rusch, Jeannie Shoveller, Chanson J Brumme, and P Richard Harrigan. Sociodemographic correlates of hiv drug resistance and access to drug resistance testing in british columbia, canada. PloS one, 12(9):e0184848, 2017.

[53] Magnus Unemo, Daniel Golparian, and David W Eyre. Antimicrobial resistance in neisseria gonorrhoeae and treatment of gonorrhea. In Neisseria gonorrhoeae, pages 37–58. Springer, 2019.

[54] Feng Fu, Daniel I Rosenbloom, Long Wang, and Martin A Nowak. Imitation dynamics of vaccination behaviour on social networks. Proceedings of the Royal Society B: Biological Sciences, 278(1702):42–49, 2010.

[55] Martina Morris and Mirjam Kretzschmar. Concurrent partnerships and the spread of hiv. Aids, 11(5):641–648, 1997.

[56] C Bauch and DA Rand. A moment closure model for sexually transmitted disease transmission through a concurrent partnership network. Proceedings of the Royal Society of London. Series B: Biological Sciences, 267(1456):2019–2027, 2000.

[57] CT Bauch. A versatile ode approximation to a network model for the spread of sexually transmitted diseases. Journal of mathematical biology, 45(5):375–395, 2002.

[58] Bruno Buonomo, Giuseppe Carbone, and Alberto d’Onofrio. Effect of seasonality on the dynamics of an imitation-based vaccination model with public health intervention. Mathematical Biosciences & Engineering, 15(1):299–321, 2018.

[59] Martial L Ndeffo Mbah, Jingzhou Liu, Chris T Bauch, Yonas I Tekel, Jan Medlock, Lauren Ancel Meyers, and Alison P Galvani. The impact of imitation on vaccination behavior in social contact networks. PLoS computational biology, 8(4):e1002469, 2012.

[60] Chris T Bauch. Imitation dynamics predict vaccinating behaviour. Proceedings of the Royal Society B: Biological Sciences, 272(1573):1669–1675, 2005.

[61] Ken TD Eames and Matt J Keeling. Modeling dynamic and network heterogeneities in the spread of sexually transmitted diseases. Proceedings of the national academy of sciences, 99(20):13330–13335, 2002.

[62] Preeti Pathela, Sarah L Braunstein, Susan Blank, and Julia A Schillinger. Hiv incidence among men with and those without sexually transmitted rectal infections: estimates from matching against an hiv case registry. Clinical infectious diseases, 57(8):1203–1209, 2013.

[63] Jason W Mitchell and Keith J Horvath. Factors associated with regular hiv testing among a sample of us msm with hiv-negative main partners. Journal of acquired immune deficiency syndromes (1999), 64(4):417, 2013.

[64] Paul A Shuper, Narges Joharchi, Hyacinth Irving, David Fletcher, Colin Kovacs, Mona Loutfy, Sharon L Walmsley, David KH Wong, and Jürgen Rehm. Differential predictors of art adherence among hiv-monoinfected versus hiv/hcv-coinfected individuals. AIDS care, 28(8):954–962, 2016.

[65] S Ha, L Pogany, J Seto, J Wu, and M Gale-Rowe. Sexually transmitted infections: What are canadian primary care physicians prescribing for the treatment of gonorrhea? Canada Communicable Disease Report, 43(2):33, 2017.

[66] Ameeta E Singh, Jennifer Gratrix, Irene Martin, Dara S Friedman, Linda Hoang, Richard Lester, Gila Metz, Gina Ogilvie, Ron Read, and Tom Wong. Gonorrhea treatment failures with oral and injectable expanded spectrum cephalosporin monotherapy vs dual therapy at 4 canadian sexually transmitted infection clinics, 2010–2013. Sexually transmitted diseases, 42(6):331–336, 2015.

